# Network analysis identifies DAPK3 as a potential biomarker for lymphovascular invasion and prognosis of colon adenocarcinoma

**DOI:** 10.1101/2021.02.03.21251102

**Authors:** Huey-Miin Chen, Justin A. MacDonald

**Author notes:** **Correspondence:** Justin MacDonald, Department of Biochemistry & Molecular Biology, University of Calgary, 3280 Hospital Drive NW, Calgary, AB, T2N 4Z6.

## Abstract

Adenocarcinoma of the colon is the fourth most common malignancy worldwide with significant rates of mortality. Hence, the identification of novel molecular biomarkers with prognostic significance is of particular importance for improvements in treatment and patient outcome. Clinical traits and RNA-Seq data of 551 patient samples and 18,205 genes in the UCSC Toil Recompute Compendium of TCGA TARGET and GTEx datasets (restricted to |Primary_site| = colon) were obtained from the Xena platform. Weighted gene co-expression network analysis was completed, and 24 unique modules were assembled to specifically examine the association between gene networks and cancer cell invasion. One module, containing 151 genes, was significantly correlated with lymphatic invasion, a histopathological feature of higher-risk colon cancer. Search tool for the retrieval of interacting genes/proteins (STRING) and gene ontology (GO) analyses identified the module to be enriched in genes related to cytoskeletal organization and apoptotic signaling, suggesting involvement in tumor cell survival and migration along with epithelial-mesenchymal transformation. Of genes that were differentially expressed and significant for overall survival, *DAPK3* (death-associated protein kinase 3) was revealed as the pseudo-hub of the module. Although *DAPK3* expression was reduced in colon cancer patients, survival analysis revealed that high expression of *DAPK3* was significantly correlated with greater lymphovascular invasion and poor overall survival.

## INTRODUCTION

Adenocarcinomas of the colon and rectum, collectively referred to as colorectal cancers (CRC), are highly aggressive and common contributors to global cancer morbidity and mortality (Dekker et al. 2019; Malki et al. 2020; Marmol et al. 2017). The likelihood of CRC diagnosis is about 5%, and risk factors include age and sex as well as diet and other lifestyle habits (Marmol et al. 2017). The distinction between colon and rectal cancer is largely anatomical, but the tumor location does impact both surgical and therapeutic management strategies as well as prognosis. In 2018, adenocarcinoma of the colon was the fourth most common malignancy and was documented to account for >10% of all diagnosed cancers globally (Ferlay et al. 2019). Colorectal adenocarcinomas are caused by genomic instability and mutations that occur in tumor suppressor genes, oncogenes, and genes related to DNA repair. Genetic and epigenetic alterations contribute to molecular events that lead to the neoplastic transformation of healthy colorectal epithelium that then progresses to malignancy (Fearon and Vogelstein 1990). Several canonical signaling pathways are known to be affected by genomic aberrations in CRC (i.e., MAPK, PI3K, TGF-β, TP53 and WNT/β-catenin), and ultimately lead to changes in fundamental cellular processes that drive tumor development, including differentiation, apoptosis, cell cycle, cell proliferation and survival (Malki et al. 2020; Marmol et al. 2017).

In addition, cancer cell migration is particularly important to the process of metastasis, in which cancer cells escape from the primary tumor and travel to distant sites to invade a new niche (Chow, Factor, and Ullman 2012). Two important pathologic processes for cancer cell migration include lymphovascular and perineural invasion, whereby tumor cells invade nervous tissue (perinuclear invasion, PNI) or small lymphatic and vascular tissues (lymphovascular invasion, LVI), respectively. Patients with locally advanced rectal cancer who demonstrated PNI and/or LVI had significantly higher risk of recurrence and poorer survival outcomes (Sun et al. 2019). Moreover, LVI and PNI had detrimental effects on survival after diagnosis of stage II or III adenocarcinoma of the colon (Mutabdzic et al. 2019; Skancke et al. 2019). In patients with stage III colon cancer, increased lymph node involvement was also associated with a need for adjuvant therapy with worsening overall survival and shorter time to recurrence (Sjo et al. 2012). An improved understanding of the genes associated with PNI and/or LNI of colon adenocarcinoma may reveal candidate biomarkers and novel therapeutic opportunities by identifying predictive and/or prognosis markers for metastatic invasion events.

In this study, we assess whether tumor molecular markers associated with colon cancer exist to improve prediction of cancer cell migration and invasion with links to overall survival. Weighted gene co-expression network analysis (WGCNA) is commonly employed to explore next generation RNA-Seq datasets for correlations among genes and clinical phenotypes (Langfelder and Horvath 2008; Li et al. 2018; Zhang and Horvath 2005). WGCNA can provide insight into the molecular networks that connect clinical features of tumor progression and has been previously used to identify candidate prognostic biomarkers or therapeutic targets (Chen et al. 2019; Zou and Jing 2019; Qin et al. 2019; Zhai et al. 2017). A co-expression module was constructed using normalized RNA-Seq data and clinicopathological outcomes for patients with colon adenocarcinoma. Search tool for the retrieval of interacting genes/proteins (STRING) network analysis and gene ontology (GO) enrichment analysis were performed to identify hub genes and the primary biological function of selected modules that were aligned with LVI and PNI.

## METHODS

### Data Retrieval

The workflow used for the study is provided in Figure 1. RNA sequencing data (RNA-Seq) for the Genotype Tissue Expression project (GTEx; https://gtexportal.org/home/) and The Cancer Genome Atlas (TCGA; https://portal.gdc.cancer.gov) were retrieved from the Xena Toil RNA-Seq Recompute Compendium (https://toil.xenahubs.net) on 30 November 2020. Although the TCGA includes normal samples, these “solid tissue normal” samples are derived from normal tissues located proximal to the tumor. Consequently, they may also possess tumor transcriptomic profiles. In contrast, samples from the GTEx project provide expression data from the normal tissue of healthy, cancer-free individuals. The datasets from the two projects cannot be compared directly, so the ‘TCGA TARGET GTEx study’ with samples restricted to |Primary_site| = ‘colon’ was retrieved from the Xena toil data hub. The Toil Compendium provides recomputed TCGA and GTEx expression raw data based on a uniform bioinformatic/RNA-Seq pipeline to eliminate batch effect due to different computational processing (Vivian et al. 2017). The Xena Toil pipeline utilized STAR for uniform realignment (Dobin et al. 2013) and applied upper-quartile normalization to RSEM quantified GTEx and TCGA gene expression (Li and Dewey 2011). The realignment also enabled the removal of degraded samples pre-quantification and batch effect correction post-quantification. The data were then filtered to exclude non-protein-coding genes. The TARGET dataset was excluded to permit focus on adult samples. Data from metastatic and recurrent tumors were also excluded due to small sample sizes. TCGA survival data and COAD clinicalMatrix information of colon cancer patients were retrieved from the Xena TCGA data hub (https://tcga.xenahubs.net). The Xena browser (http://xena.ucsc.edu), hosted by the Computational Genomics Lab of the University of California Santa Cruz (UCSC; https://cglgenomics.ucsc.edu), was utilized to compare gene expressions (Goldman et al. 2020). Data processing was completed using the R (v4.0.2) programming language, and data retrieval was completed via the UCSCXenaTools R package.

**Figure 1.**
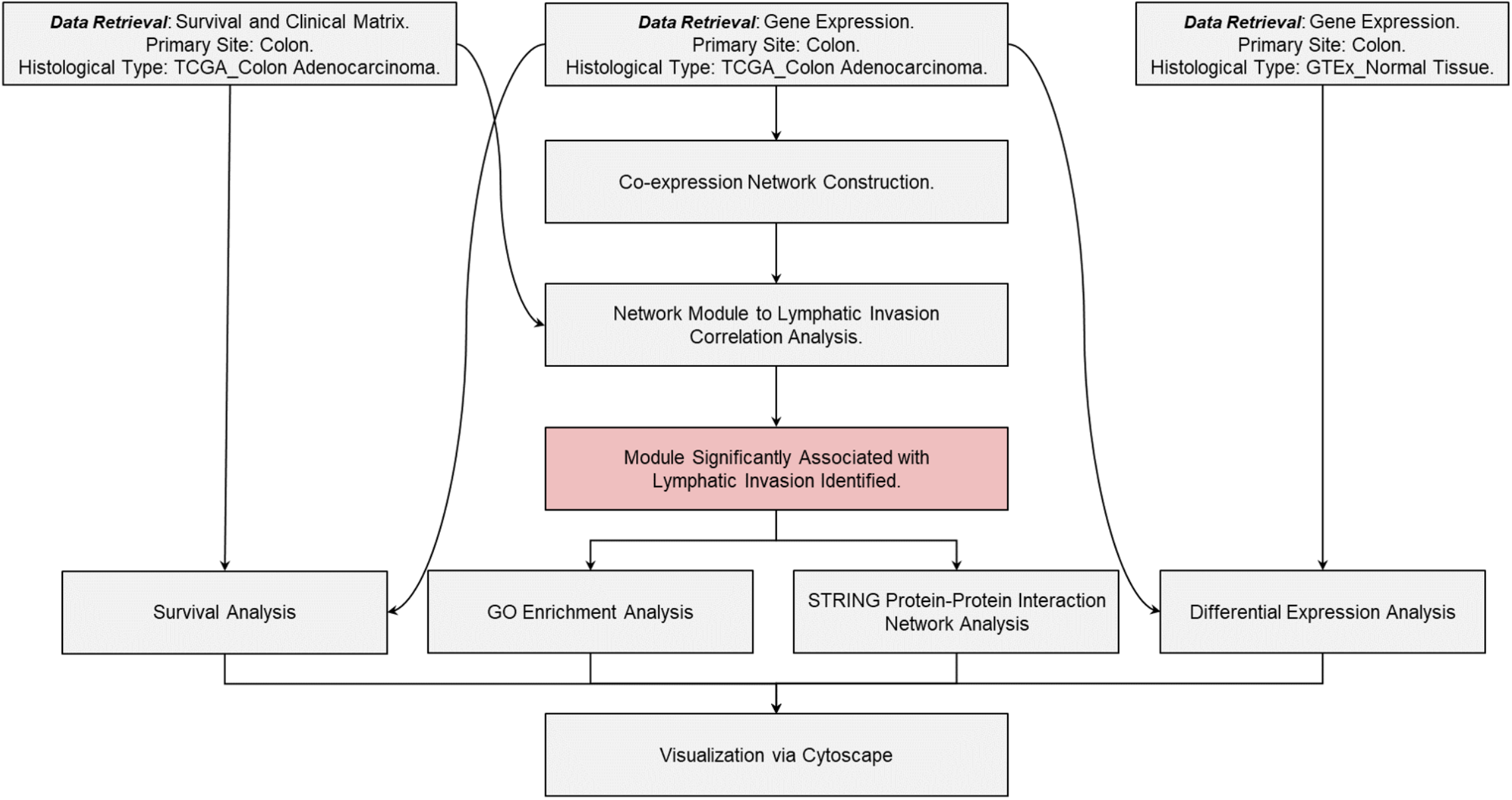
Workflow Scheme. The workflow for data acquisition, preparation, processing and analyses.

### Data Pre-processing

The log-transformed expected count retrieved from Xena Toil was back-transformed via the |round(((2^x)-1),0)| function. The data were further refined to include only ‘Normal Tissue’ from the GTEx gene sets and only ‘Primary Tumor’ with |histological_type| = ‘Colon Adenocarcinoma’ from the TCGA gene sets. After these pre-processing steps, the dataset included 551 patient samples and 18,205 genes.

### Identification of Differentially Expressed Genes

The edgeR-limma workflow was used to detect differentially expressed genes (DEG) between cancer and normal samples (Law et al. 2016). The input data (18,205 genes, 551 samples) were screened with function |filterByExpr.default| to eliminate outliers and lowly-expressed genes. The final DGEList object contained 15,164 genes and 547 sample (303 GTEx, 244 TCGA). Heteroscedasticity was removed via function |voom|. Expression values of TCGA were then compared to that of GTEx through linear modelling; namely, the limma functions |lmFit|, |contrasts.fit|, and |eBayes|. All functions were operated with default settings. The transformed expression values, generated by |voom|, were exported for subsequent analyses

### Weighted gene co-expression network analysis (WGCNA)

To identify candidate biomarkers for lymphatic invasion within the colon adenocarcinoma subset, we applied WGCNA to voom-transformed expression values (Langfelder and Horvath 2008). The network topology was calculated for a range of soft-thresholding power (β). Then, the scale-free topology fit index was plotted as a function of the soft-thresholding power, and the mean connectivity as a function of the soft-thresholding power was determined. Although β = 9 was the lowest power for which the scale-free topology fit index reached an R^2^ value of 0.90, β = 10 was ultimately used as it was recommended as the minimum soft-threshold power to be employed for a signed network (Langfelder and Horvath 2008). The co-expression similarity was raised to achieve scale-free topology with this soft-thresholding power. Co-expression similarity and adjacency were calculated for a signed network, transformed into a topological overlap matrix (TOM) to calculate the corresponding dissimilarity (dissTOM) which in turn was used to conduct hierarchical clustering. Module identification was completed with dynamic tree cut, and any highly similar modules with the height of clustering lower than 0.25 were merged. A clustering dendrogram was used to display the results.

### Generation of module-trait relationships and pathway enrichment analysis of genes

Correlations between modules generated from the WGCNA and clinical parameters were determined by module-trait relationship (MTR) analysis. Module eigengenes (MEs) were calculated and related to clinical parameters. Gene ontology (GO) enrichment analysis was completed with topGO R to facilitate biological interpretation. Finally, intramodular connectivity was analyzed using the stringApp (Doncheva et al. 2019; Szklarczyk et al. 2021) and visualized with Cytoscape v3.8.2 (Otasek et al. 2019; Shannon et al. 2003).

### Survival Analysis

Survival data were retrieved as part of the TCGA dataset downloaded from Xena and were subjected to Kaplan-Meier analysis in R. The Cox proportional-hazards model was used to investigate the association between survival time and candidate biomarkers for lymphatic invasion. Z-scale cut-offs were set at *≥* 0.647 for high expression and *≤* 0.647 for low expression. Survival time cut off was set at 10 years. The coxph R function was utilized in combination with the |log-rank| test for comparison of the high/low survival curves. Survival curves were plotted to show the differences in patient mortality between high- and low-expression groups.

### Differential gene correlation analysis (DGCA)

Using the R package DGCA (McKenzie et al. 2016), the differential correlation between *DAPK3* and a filtered list of human protein-coding genes (8,528) was computed with the |ddCorAll| function. This pipeline provided the Pearson coefficient (*r*) and the corresponding *p* values for each pair of genes across samples (n=547). Significant changes in differential correlation between the two conditions (healthy colon vs. colon adenocarcinoma) were then identified using a Fisher’s *Z*-test.

## RESULTS

The presence of tumor cells within the lymphatics or blood vessels is associated with increased risk of axillary lymph node and distant metastases. To identify candidate biomarkers for lymphatic invasion, we performed WGCNA on RNA-Seq data obtained from TCGA with the |primary_site| originating from colon, |histological_type| limited to colon adenocarcinoma, and non-protein-coding genes excluded. When the soft thresholding was set at ten, the scale-free topology fit index reached 0.95 (Figure 2A, 2B). Modules of co-expressed genes were determined with hierarchical clustering and the dynamic tree cut procedure, and each of the modules was marked by a color. In all, 24 different modules resulted from WGCNA with the clustering dendrograms of genes shown in Figure 2C. Merging of modules with similar expression profiles was completed with a cut height of 0.25, corresponding to a 0.75 correlation; however, this did not reduce the number of modules. The *darkgrey* module was the smallest, with 100 genes, while the *turquoise* and *blue* modules were the largest, with 2,005 and 1,742 genes, respectively (Table 1). All genes were assigned, and no genes were grouped in *grey* (i.e., the module reserved for genes with no assigned classification).

**Table 1.** WGCNA-derived gene modules, gene count of membership, and associated GO terms. GO terms were deemed significant if |p value| ≤ 0.01 and |total number of annotated genes| ≥ 30. GO terms are listed by odds ratio.

| Module | #Genes | Enriched GO Terms | Module | #Genes | Enriched GO Terms |
| --- | --- | --- | --- | --- | --- |
| turquoise | 2,005 | GO:0006119:oxidative phosphorylation<br>GO:0022904:respiratory electron transport chain<br>GO:0033108:mitochondrial respiratory chain complex...<br>GO:0032543:mitochondrial translation | salmon | 225 | GO:0019674:NAD metabolic process<br>GO:0034198:cellular response to amino acid starvati...<br>GO:0006734:NADH metabolic process<br>GO:0061337:cardiac conduction |
| blue | 1,742 | GO:0035904:aorta development<br>GO:0030500:regulation of bone mineralization<br>GO:0045123:cellular extravasation<br>GO:0006029:proteoglycan metabolic process | cyan | 185 | GO:0006378:mRNA polyadenylation<br>GO:0000724:double-strand break repair via homologou... |
| brown | 811 | GO:0002011:morphogenesis of an epithelial sheet<br>GO:0045652:regulation of megakaryocyte differentiat...<br>GO:0051898:negative regulation of protein kinase B ...<br>GO:0038202:TORC1 signaling | midnightblue | 180 | GO:0006397:mRNA processing<br>GO:0000398:mRNA splicing, via spliceosome<br>GO:0031331:positive regulation of cellular cataboli...<br>GO:2000113:negative regulation of cellular macromol... |
| yellow | 644 | GO:0016266:O-glycan processing<br>GO:1902476:chloride transmembrane transport<br>GO:0050891:multicellular organismal water homeostas...<br>GO:0150104:transport across blood-brain barrier | lightcyan | 171 | GO:1901992:positive regulation of mitotic cell cycl...<br>GO:1990542:mitochondrial transmembrane transport |
| green | 579 | GO:0006614:SRP-dependent cotranslational protein ta...<br>GO:0000184:nuclear-transcribed mRNA catabolic proce...<br>GO:0006413:translational initiation<br>GO:0019083:viral transcription | grey60 | 162 | GO:0060324:face development<br>GO:1900006:positive regulation of dendrite developm...<br>GO:0048008:platelet-derived growth factor receptor ...<br>GO:0030510:regulation of BMP signaling pathway |
| red | 441 | GO:0033260:nuclear DNA replication<br>GO:2000816:negative regulation of mitotic sister ch...<br>GO:0030071:regulation of mitotic metaphase/anaphase...<br>GO:0140013:meiotic nuclear division | lightgreen | 157 | GO:0030433:ubiquitin-dependent ERAD pathway<br>GO:1900542:regulation of purine nucleotide metaboli...<br>GO:0006366:transcription by RNA polymerase II |
| black | 347 | GO:0046580:negative regulation of Ras protein signa...<br>GO:0035825:homologous recombination<br>GO:0006397:mRNA processing<br>GO:0043087:regulation of GTPase activity | lightyellow | 156 | GO:0090329:regulation of DNA-dependent DNA replicat...<br>GO:0071364:cellular response to epidermal growth fa...<br>GO:0042147:retrograde transport, endosome to Golgi<br>GO:0008203:cholesterol metabolic process |
| pink | 322 | GO:0032509:endosome transport via multivesicular bo...<br>GO:0007099:centriole replication<br>GO:0051972:regulation of telomerase activity<br>GO:0007080:mitotic metaphase plate congression | royalblue | 153 | GO:0009247:glycolipid biosynthetic process<br>GO:0009566:fertilization<br>GO:0090181:regulation of cholesterol metabolic proc... |
| magenta | 297 | GO:0006521:regulation of cellular amino acid metabo...<br>GO:0090151:establishment of protein localization to...<br>GO:1902036:regulation of hematopoietic stem cell di...<br>GO:0090502:RNA phosphodiester bond hydrolysis, endo... | darkred | 151 | GO:1902108:regulation of mitochondrial membrane per...<br>GO:2001235:positive regulation of apoptotic signali...<br>GO:0010821:regulation of mitochondrion organization<br>GO:0030036:actin cytoskeleton organization |
| purple | 238 | GO:0006378:mRNA polyadenylation<br>GO:0006369:termination of RNA polymerase II transcr...<br>GO:0009303:rRNA transcription<br>GO:0006409:tRNA export from nucleus | darkgreen | 114 | GO:0034314:Arp2/3 complex-mediated actin nucleation<br>GO:0019068:virion assembly<br>GO:0006890:retrograde vesicle-mediated transport, G...<br>GO:0010970:transport along microtubule |
| greenyellow | 235 | GO:0032365:intracellular lipid transport<br>GO:0044088:regulation of vacuole organization<br>GO:0006635:fatty acid beta-oxidation<br>GO:0015850:organic hydroxy compound transport | darkturquoise | 101 | GO:1903146:regulation of autophagy of mitochondrion<br>GO:0006890:retrograde vesicle-mediated transport, G...<br>GO:0043666:regulation of phosphoprotein phosphatase...<br>GO:0010822:positive regulation of mitochondrion org... |
| Tan | 234 | GO:0070423:nucleotide-binding oligomerization domai...<br>GO:0099072:regulation of postsynaptic membrane neur...<br>GO:0006826:iron ion transport<br>GO:1902600:proton transmembrane transport | darkgrey | 100 | None. |

**Figure 2.**
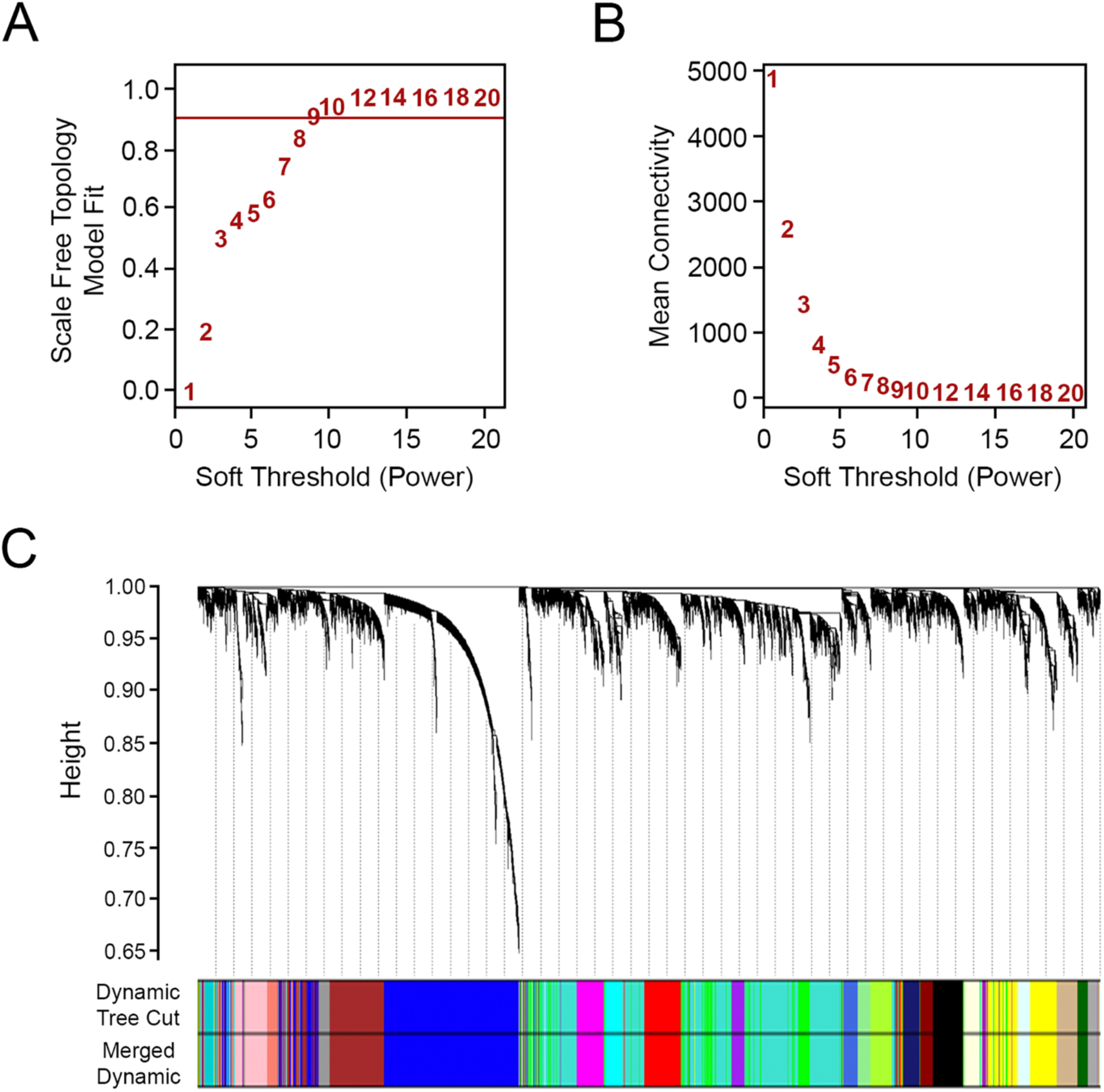
Weighted gene co-expression network analysis of primary tumors from colon adenocarcinoma. A range of soft thresholding power (β) was screened within the WGCNA pipeline (**A**), and the plot shows the scale-free topology fit index for different β powers. The mean connectivity for various soft thresholding powers is also provided (**B**). Hierarchical clustering and module assignment was completed based on a topological overlap matrix to calculate the corresponding dissimilarity (dissTOM). In (**C**), the connected genes were grouped into colored modules with the dynamic tree cut method. Highly correlated modules were merged based on a cut height of 0.25.

The clinical parameters of colon adenocarcinoma patients, including lymphatic invasion, venous invasion and perineural invasion occurrence as well as pathological stage and pathological T (tumor, T), pathological N (node, N) and pathological M (metastasis, M) staging categories were extracted from the TCGA dataset and then related to the module eigengenes (ME). As shown in Figure 3, ME*darkred* was correlated with lymphatic invasion (r = 0.13, p = 0.04) and venous invasion (r = 0.14, p = 0.04). Additional correlations were observed for ME*darkred* with pathology N (r = 0.14, p = 0.03) and pathological stage (r = 0.17, p =0.01). Notably, the *darkred* module contained *DAPK3* (death-associated protein kinase 3) along with 150 other genes (Table 2). *DAPK3* expression is attenuated in several squamous cell carcinomas (Kake et al. 2017; Li et al. 2015; Kocher, White, and Piwnica-Worms 2015; Bi et al. 2009; Das et al. 2016). Moreover, loss-of-function mutations or deletion of *DAPK3* promote increased cell survival, proliferation, cellular aggregation, and increased resistance to chemotherapy (Brognard et al. 2011). In light of this evidence, *DAPK3* is now strongly implicated as a tumor suppressing kinase.

**Table 2.** Gene expression log*2*FC and hazard ratio (HR, high vs. low expression) of genes in the *darkred* module. Significant values are highlighted in green; For differential expression, significance was identified with |log_2_FC| > 0.58 AND |FDR| ≤ 0.05. For HR, significance was identified with |likelihood ratio test value| ≤ 0.05.

| Gene | Log<sub>2</sub>FC | HR | Gene | Log<sub>2</sub>FC | HR | Gene | Log<sub>2</sub>FC | HR |
| --- | --- | --- | --- | --- | --- | --- | --- | --- |
| *ENKD1* | -1.21 | 3.63 | *INO80E* | -1.28 | 1.81 | *ELMO3* | 2.54 | 1.30 |
| *CDIPT* | -0.57 | 3.61 | *RNPEPL1* | 0.00 | 1.62 | *LLGL2* | 2.76 | 0.79 |
| *NOL3* | -0.84 | 3.04 | *NSMF* | 0.13 | 1.65 | *SMPD4* | 0.28 | 1.26 |
| *ACD* | 0.09 | 2.83 | *COG4* | 0.07 | 1.72 | *FAM160A2* | -0.75 | 0.79 |
| *MAN1B1* | 0.08 | 2.52 | *NOXA1* | -0.20 | 1.58 | *FLYWCH1* | -1.51 | 1.29 |
| *LIME1* | 0.03 | 2.60 | *C16orf58* | 0.05 | 1.64 | *PIDD1* | -1.22 | 1.30 |
| *ARRDC1* | 0.80 | 2.60 | *COPS7B* | -0.32 | 1.59 | *SETD6* | -0.23 | 1.24 |
| *VAC14* | 0.48 | 2.51 | *SPNS1* | -0.01 | 1.68 | *BET1L* | 0.05 | 1.26 |
| *WDR24* | -0.14 | 2.40 | *ZNF48* | 0.25 | 1.66 | *HAUS5* | 0.19 | 1.20 |
| *CTDSP1* | -0.29 | 2.84 | *SPATA33* | 0.63 | 1.62 | *EPS8L2* | 1.01 | 0.83 |
| *TUT1* | 0.07 | 2.51 | *ACSF3* | 0.06 | 1.57 | *TELO2* | 0.22 | 0.82 |
| *NPRL3* | 0.60 | 2.61 | *ZNF768* | -0.21 | 1.59 | *GALT* | -1.14 | 1.21 |
| *ANKRD13D* | -0.95 | 2.28 | *MAP3K11* | -0.03 | 1.63 | *PIGQ* | -0.34 | 1.18 |
| *DHODH* | 0.41 | 2.39 | *SIGIRR* | -0.10 | 1.56 | *RABEP2* | 0.31 | 1.16 |
| *DEF8* | -0.65 | 2.33 | *TRAF2* | 0.95 | 1.52 | *AXIN1* | 0.59 | 0.87 |
| *CRACR2B* | 0.61 | 2.08 | *ZNF205* | 0.10 | 1.53 | *CAPN15* | 0.40 | 0.86 |
| *FHOD1* | -0.54 | 2.05 | *SSH3* | -0.15 | 1.54 | *CTTN* | -0.20 | 1.16 |
| *STK25* | -0.48 | 2.12 | *SHROOM1* | 0.20 | 0.67 | *MXD3* | -0.73 | 0.86 |
| *DAPK3* | -0.98 | 2.05 | *LRSAM1* | -0.56 | 1.51 | *FUK* | 0.17 | 1.14 |
| *ASB6* | -0.15 | 2.06 | *EIF2B5* | -0.36 | 1.67 | *DIS3L2* | 0.01 | 1.12 |
| *VPS4A* | -0.11 | 2.03 | *MICALL2* | -1.29 | 1.55 | *TMEM8A* | 0.71 | 0.86 |
| *B3GALT6* | 0.62 | 2.10 | *TMCO6* | -0.33 | 1.50 | *ESRRA* | 0.60 | 0.88 |
| *PARP10* | -0.49 | 2.05 | *CD2BP2* | -0.08 | 1.45 | *KLHL17* | -1.40 | 1.13 |
| *UBALD1* | -0.83 | 1.99 | *RTKN* | 1.32 | 1.49 | *ACAP3* | -2.07 | 1.14 |
| *BANP* | -0.21 | 2.08 | *STX4* | -0.27 | 1.55 | *WDR90* | -0.28 | 0.89 |
| *ZNF668* | 0.37 | 2.01 | *E4F1* | -0.77 | 1.49 | *CCHCR1* | 0.16 | 1.12 |
| *ARMC5* | -0.40 | 1.98 | *MUTYH* | -0.05 | 1.49 | *RHOT2* | -1.10 | 1.11 |
| *KAT8* | -0.63 | 2.03 | *WDR59* | -0.44 | 1.47 | *C2CD2L* | -0.54 | 0.91 |
| *GPR35* | 2.51 | 0.51 | *RAB17* | 2.10 | 1.48 | *CD151* | 0.05 | 1.10 |
| *AMDHD2* | -0.96 | 1.97 | *PRR14* | -0.54 | 1.48 | *RAPGEFL1* | 1.05 | 0.91 |
| *RAD9A* | -0.43 | 2.13 | *CCDC24* | 0.09 | 1.39 | *TOLLIP* | -0.01 | 0.91 |
| *PARN* | 0.39 | 1.86 | *PLEKHH3* | -1.08 | 1.48 | *HIP1R* | 0.79 | 0.91 |
| *CDK9* | -0.86 | 1.88 | *SLC27A4* | 0.56 | 0.68 | *PWWP2B* | -0.06 | 1.08 |
| *GAS8* | -0.24 | 1.88 | *TFAP4* | 1.64 | 1.41 | *ARHGAP27* | 1.00 | 0.91 |
| *SPSB3* | -1.34 | 1.83 | *MLYCD* | -0.59 | 0.72 | *KCNQ1* | 3.82 | 0.92 |
| *PAQR4* | 2.91 | 1.94 | *MYO7B* | 0.95 | 0.67 | *SH2B1* | -1.38 | 1.09 |
| *SLC38A7* | 0.35 | 1.73 | *ABCF3* | 0.09 | 1.38 | *SIRT3* | -0.58 | 1.07 |
| *RHBDF1* | -0.52 | 1.89 | *PNPLA2* | -1.07 | 1.40 | *SGSH* | -0.66 | 0.92 |
| *E2F4* | 0.18 | 1.79 | *TRAF7* | 0.49 | 1.37 | *KATNB1* | 0.80 | 0.93 |
| *NAPRT* | 0.70 | 2.05 | *CORO7* | 0.34 | 0.72 | *DDX51* | -0.46 | 1.06 |
| *MOB2* | -1.41 | 1.80 | *TPRN* | 1.56 | 0.75 | *ZNF213* | -0.01 | 1.06 |
| *ING5* | -0.65 | 1.76 | *ROGDI* | -1.94 | 1.36 | *FBXO31* | -0.87 | 0.95 |
| *RIPK4* | 2.13 | 1.90 | *PHKG2* | -0.34 | 1.35 | *FBXW5* | -0.68 | 0.96 |
| *ATG4B* | -1.06 | 1.75 | *CC2D1A* | 0.03 | 0.74 | *NADSYN1* | -0.46 | 1.03 |
| *MGAT4B* | 0.95 | 0.57 | *TBC1D24* | -0.06 | 1.29 | *SH3GLB2* | -0.88 | 0.97 |
| *CHTF18* | 0.60 | 1.77 | *RHPN1* | 0.84 | 1.34 | *ESRP2* | 3.51 | 1.03 |
| *PRMT7* | -0.10 | 1.89 | *RASSF7* | 2.20 | 1.33 | *ZNF598* | 0.21 | 0.97 |
| *NELFB* | 0.34 | 1.85 | *CDK11B* | -0.57 | 0.74 | *TMEM234* | -0.68 | 0.97 |
| *TBC1D10B* | 0.33 | 1.83 | *NAA60* | -0.04 | 1.31 | *SIRT7* | -0.36 | 0.97 |
| *MOGS* | 0.34 | 1.75 | *EXD3* | -1.34 | 1.26 |  |  |  |
| *TESK1* | -0.82 | 1.71 | *RAB40C* | -0.13 | 1.27 |  |  |  |

**Table 3.** Hub gene prioritization by network analysis of protein-protein interaction. Hub and pseudohub gene(s) were selected base on *degree distribution* and *closeness centrality*. Highlighted are DEGs that significantly impacted overall survival probability.

| Gene | Degree<sup>1</sup> | Closeness Centrality<sup>2</sup> | Neighbourhood Connectivity<sup>3</sup> | Radiality<sup>4</sup> | Stress<sup>5</sup> | Eccentricity<sup>6</sup> | Betweenness Centrality<sup>7</sup> |
| --- | --- | --- | --- | --- | --- | --- | --- |
| *RHOT2* | 38 | 0.545 | 13.82 | 0.978 | 13474 | 3 | 0.125 |
| *SSH3* | 33 | 0.526 | 15.85 | 0.976 | 8744 | 3 | 0.070 |
| *RIPK4* | 33 | 0.509 | 13.39 | 0.975 | 9260 | 4 | 0.077 |
| *PIDD1* | 31 | 0.507 | 14.84 | 0.974 | 6562 | 4 | 0.050 |
| *TELO2* | 25 | 0.486 | 14.76 | 0.972 | 5780 | 4 | 0.056 |
| *DAPK3* | 10 | 0.409 | 13.70 | 0.962 | 810 | 4 | 0.006 |
| *ARRDC1* | 7 | 0.401 | 16.86 | 0.961 | 666 | 4 | 0.005 |
| *NOL3* | 4 | 0.347 | 10.75 | 0.950 | 126 | 5 | 0.001 |
| *DEF8* | 3 | 0.334 | 10.67 | 0.948 | 128 | 5 | 0.001 |
| *ANKRD13D* | 1 | 0.258 | 6.00 | 0.924 | 0 | 6 | 0.000 |
| *ENKD1* | 1 | 0.269 | 9.00 | 0.929 | 0 | 6 | 0.000 |

<sup>1</sup> Degree: the number of interactions linked to a given gene.
<sup>2</sup> Closeness centrality: a measure of how fast information spreads from a given gene to other reachable genes in the network.
<sup>3</sup> Neighbourhood connectivity: the average connectivity of neighbours of a given gene.
<sup>4</sup> Radiality: a node centrality index.
<sup>5</sup> Stress: the number of shortest paths passing through a given gene.
<sup>6</sup> Eccentricity: the maximum non-infinite length of shortest path between a given gene and another gene in the network.
<sup>7</sup> Betweenness centrality: the amount of control that a given gene exerts over the interactions of other genes in the network.

**Figure 3.**
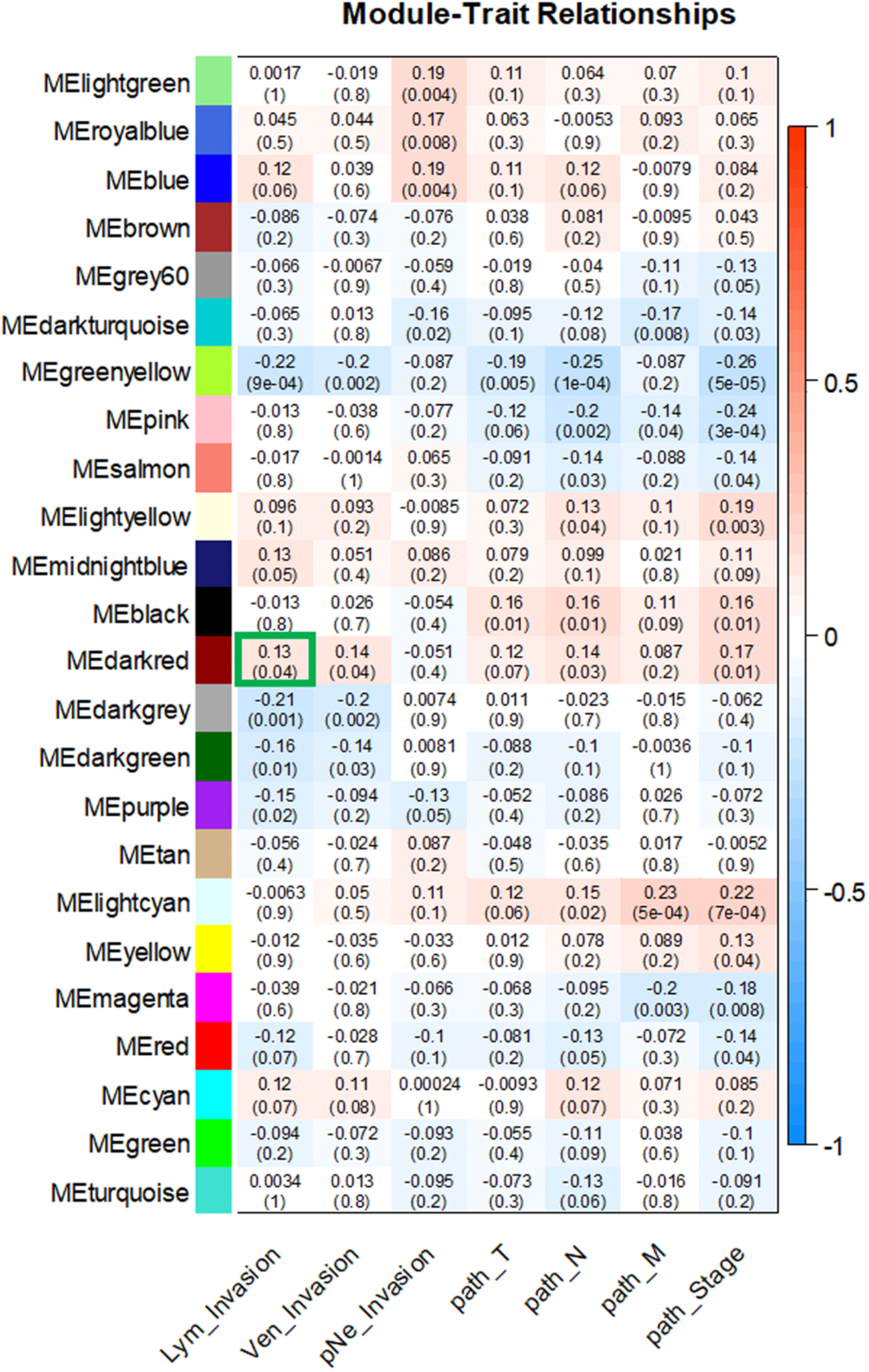
Module-trait relationship matrix correlating MEs with clinical traits. For the module-trait relationships, each row in the matrix corresponds to a module eigengene (ME) and each column to a clinical trait. Each cell contains the corresponding correlation with the p-values of the correlations in parentheses. Each module-trait relationship is color-coded by correlation as indicated in the color legend on the right; blue indicates a negative correlation while red represents a positive one. The ME*darkred* (outlined in green) had the highest correlation with lymphatic invasion and contains *DAPK3*.

To further validate the module-clinical trait relationships, we completed additional examination of the correlation between the gene significance (GS) and the module membership (MM) measures. Five modules possessed a greater proportion of gene members with positive significance towards lymphatic invasion, including the *darkred* module (r = 0.29; p = 0.0003) as well as *blue, cyan, pink*, and *lightyellow* modules (Figure 4). As the module of primary interest, *darkred* contained a majority of members (79%) with positive gene significance towards lymphatic invasion. MEs were also used as representative profiles to assess module similarity by eigengene correlation. Construction of the eigengene network provided the identification of groups of correlated eigengenes termed meta-modules. ME*darkred*, ME*black*, ME*midnightblue* and ME*lightyellow* modules were highly related. The mutual correlation of these four MEs was much stronger than their correlation with lymphatic invasion; however, lymphatic invasion was present as a part of the meta-module (Figure 5).

**Figure 4.**
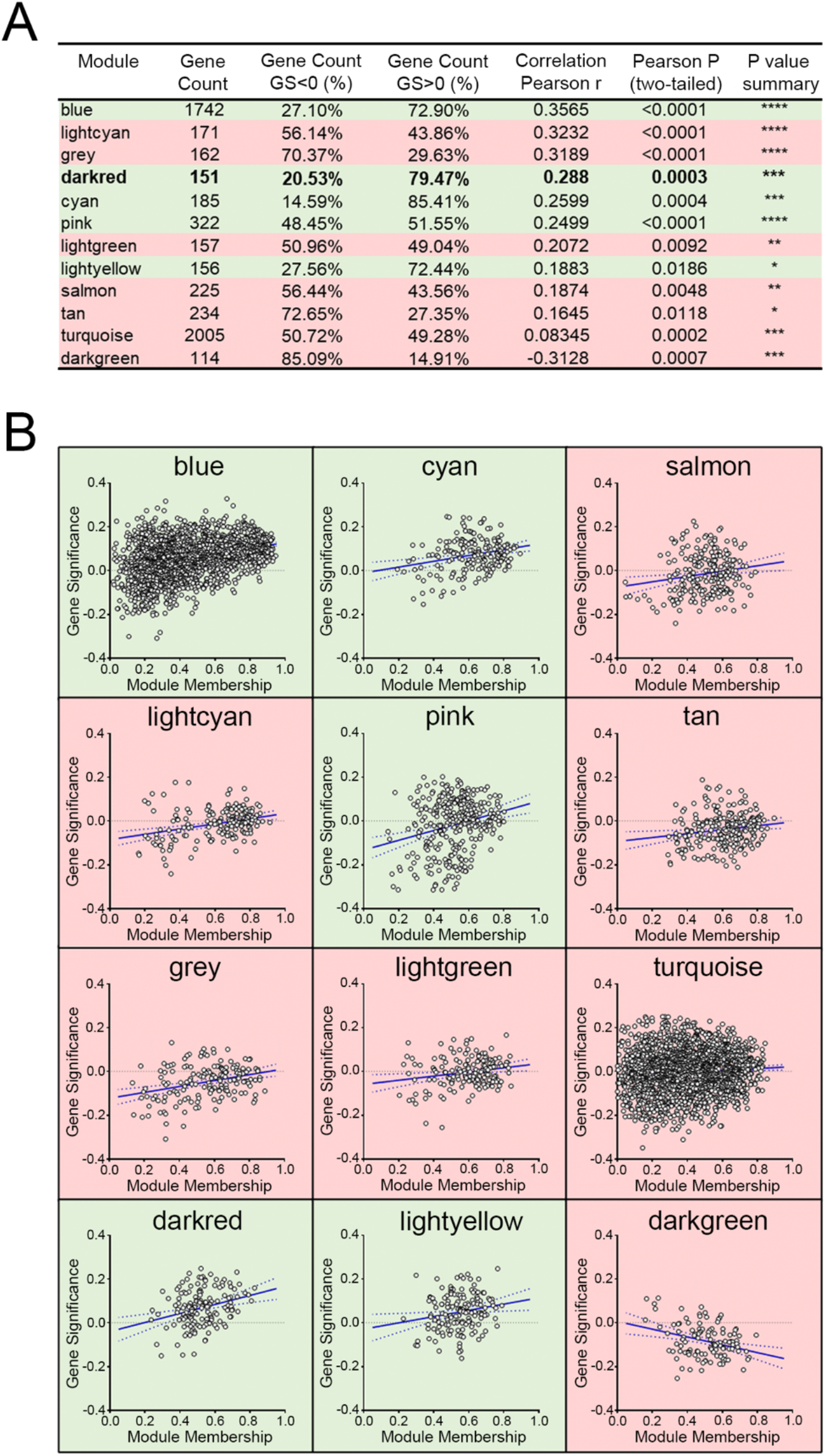
Intramodular analysis for modular membership versus gene significance. (**A**) The correlation between the gene significance (GS) and module membership (MM) measures was explored. Shown are the Pearson correlations for modules with p-values (two-tailed) indicating significance. (**B**) Scatter plots of GS versus MM for lymphatic invasion. GS is the relationship between the gene and the clinical trait, and the MM describes the correlation between the ME and the expression profile of a given gene with said module. Highlighted in green are modules with a majority of members having positive gene significance towards lymphatic invasion. Red highlights modules that possess a negative gene significance.

**Figure 5.**
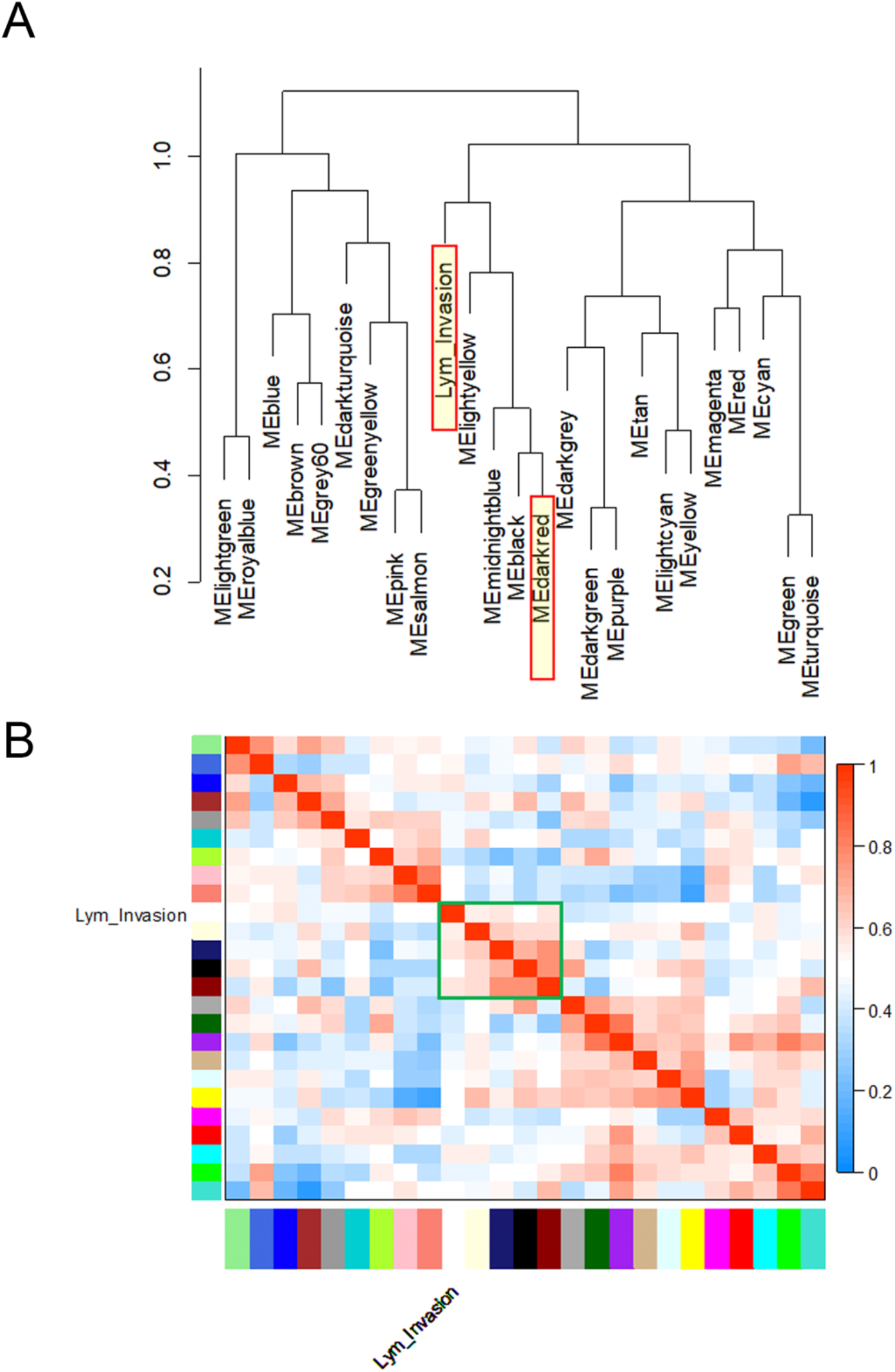
Eigengene network of meta-modules. Module eigengenes (MEs) were used as representative profiles to quantify module similarity by eigengene correlation. (**A**) Dendrogram of the MEs and the ‘lymphatic invasion’ clinical trait. (B) Heatmap of the ME-ME or ME-trait relationships. The *darkred, black, mightblue*, and *lightyellow* modules were highly related within a meta-module (outlined in green). However, the mutual correlation of individual meta-module members was stronger than their correlation with lymphatic invasion.

Differential expression analysis is routinely used to interpret the biological importance of transcriptomics experiments (Crow et al. 2019). RNA-Seq data from TCGA and GTEx were analyzed to obtain a list of differentially-expressed genes (DEGs). Based on the threshold of |log2(FC)| > 0.58 and an adjusted p-value cut-off of 5%, 9,390 DEGs were identified. For a stricter definition of significance, the treat method was used to calculate the p-values from empirical Bayes-moderated t-statistics with a minimum |log2(FC)| requirement (McCarthy and Smyth 2009). The number of DEGs was reduced to a total of 8,479 when treat testing required a |log2(FC)| greater than 0.58. Among the 8,479 DEGs, 3,579 were significantly upregulated in CAC samples whereas 4,900 were significantly downregulated in CAC samples (Figure 6A).

**Figure 6.**
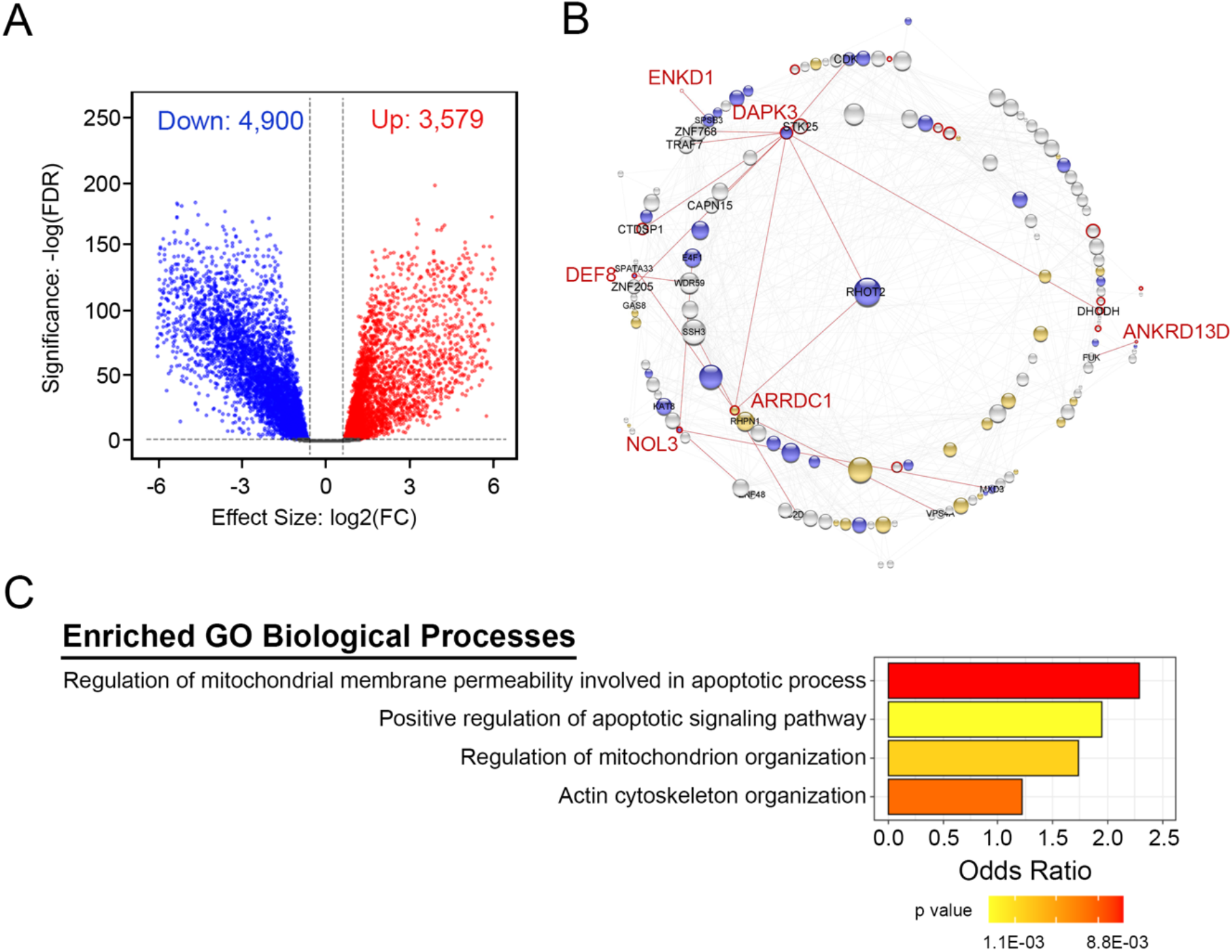
Biomolecular interaction network constructed for *DAPK3* and other gene products of the *darkred* module. A volcano plot of differentially-expressed genes (DEGs) derived from the TCGA and GTEx datasets and plotted as the log2-fold change differences (log2FC) between the compared samples (**A**). The up-regulated (red) and down-regulated (blue) genes were assessed for statistical significance using the *treat* method with empirical Bayes-moderated t-statistics (-log(FDR)). In **(B)**, genes within the *darkred* module were used as input for protein-protein interaction network construction using the search tool for the retrieval of interacting genes/proteins (STRING). The network was visualized with Cytoscape. The node size reflects the degree of interactions (i.e., edges) linked to a given gene. The node color describes the differential expression: blue, down-regulation; orange, up-regulation; and white, not differentially expressed. The node border color is reflective of the association with overall survival: red, |log_rank < 0.05| and blank, not significant. Genes that were differentially expressed and significantly associated with overall survival are noted in red text. In (**C**), gene ontology (GO) enrichment analysis was conducted for genes in the *darkred* module. Biological process (BP) GO terms are listed along the y-axis. GO terms were deemed significant when |p value| ≤ 0.01 and |total number of annotated genes| ≥ 30.

The list of DEGs generated via the limma R pipeline was amalgamated with WGCNA-derived modules. An examination of the consolidated analyses revealed 57/151 (38%) of genes in the *darkred* module to be differentially expressed. As shown in Figure 6B, genes within the *darkred* module were used as input for protein-protein interaction network construction with the STRING database. Intramodular connectivity was used to identify putative intramodular hub genes with the top five candidates having greater than 25 intramodular connections (i.e., RHOT2, mitochondrial Rho GTPase 2; RIPK3, receptor-interacting serine/threonine-protein kinase 3; SSH3, slingshot homolog 3; PIDD1, p53-induced death domain-containing protein 1; and TELO2, telomer length regulation protein TEL2 homolog). Functional enrichment analysis was conducted with topGO R on the *darkred* module, and the enriched GO biological processes were primarily found to be dominated by regulation of cellular architecture (Figure 6C). Importantly, the significantly enriched pathways were aligned with “Actin cytoskeleton organization; GO:0030036”, the “Positive regulation of apoptotic signaling pathway; GO:2001235”, the “Regulation of mitochondrion organization; GO:0010821” and the “Regulation of mitochondrial membrane permeability involved in apoptotic process; GO:1902108”.

The R function *coxph* |survival| was used to determine the association between differential gene expression and overall survival. This analysis identified 18/151 (12%) genes in the *darkred* module that passed the |log_rank = 0.05| threshold. Of this subset, 6/151 genes were significantly differentially expressed and significantly associated with overall survival (Figure 7). Importantly, none of the five hub candidates were both differentially expressed and linked to overall survival probability. Of genes that were differentially expressed and were significant for overall survival, *DAPK3* was revealed as the pseudo-hub gene of the network (Figure 6B). It also possessed a high degree of connectivity as revealed by degree distribution and closeness centrality (Supplemental Table S3). In addition to *DAPK3, ANKRD13D* (ankyrin repeat domain-containing protein 13D), *ENKD1* (enkurin domain-containing protein 1), *DEF8* (differentially expressed in FDCP 8 homolog), *NOL3* (nucleolar protein 3) and *ARRDC1* (*α*-arrestin domain-containing protein 1) exhibited differential expression profiles that significantly impacted on overall survival probability.

**Figure 7.**
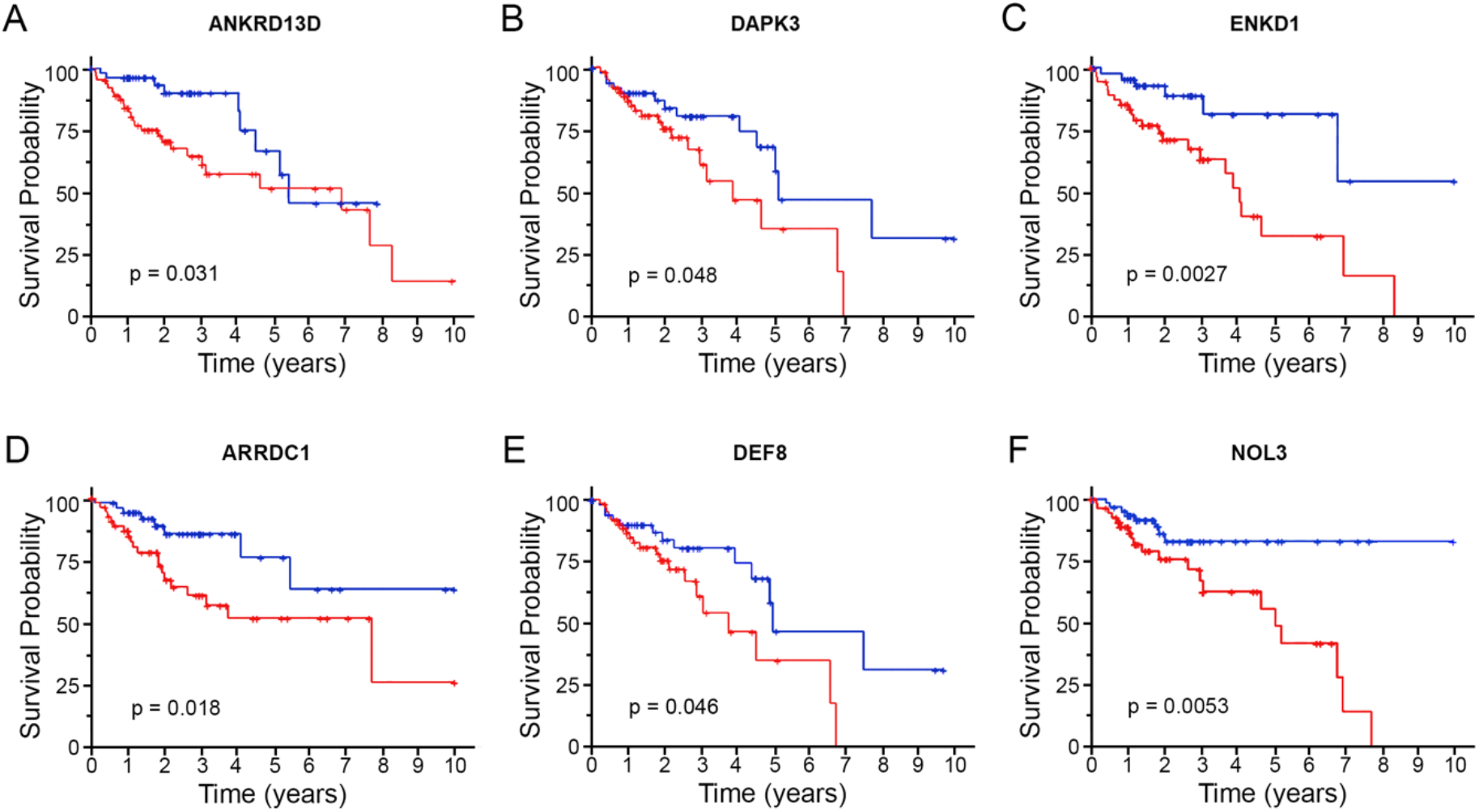
Survival analysis of consequential DEGs within the *darkred* module. Six genes were revealed to be differentially expressed and be significantly associated with overall survival. Kaplan-Meier survival analysis by log-rank test was used to examine survivability with respect to high or low expression of (**A**) *ANKRD13D*, ankyrin repeat domain-containing protein 13D; (**B**) *DAPK3*, death-associated protein kinase 3; (**C**) *ENKD1*, enkurin domain-containing protein 1; (**D**) *ARRDC1*, arrestin domain-containing protein 1; (**E**) *DEF8*, differentially expressed in FDCP 8 homolog; and (**F**) *NOL3*, nucleolar protein 3.

To investigate whether *DAPK3* was abnormally expressed in human colon cancer, the Toil recomputed expression of *DAPK3* in GTEx and TCGA (healthy colon vs. colon adenocarcinoma, primary tumor) was juxtaposed via unpaired t-test with Welch’s correction. As shown, the level of *DAPK3* expression is significantly downregulated (Figure 8A; -0.878 ± 0.048, p<0.0001) in colon adenocarcinoma. A subsequent analysis of *DAPK3* expression (healthy colon vs. colon adenocarcinoma, primary tumor; unified by (Wang et al. 2018) generated consistent results and verified reduced *DAPK3* expression in colon cancer samples (Figure 8B; -0.825 ± 0.089, p<0.0001). Thus, the gene expression analysis further substantiates *DAPK3* as a candidate tumor suppressor gene. Survival data was also retrieved as part of the UCSC dataset downloaded from Xena and subjected to Kaplan-Meier analysis to determine the significance of *DAPK3* expression on survival probability in patients with colon adenocarcinoma. Figure 9A shows that high *DAPK3* expression (>10.92, upper quartile) strongly correlates with poor overall survival probability when compared with low DAPK3 expression (<10.39, lower quartile). However, *DAPK3* expression does not make a significant impact on disease specific survival (Figure 9B). One feasible explanation for this discrepancy is an experimentally defined role for *DAPK3* in actin filament and focal adhesion dynamics (Nehru, Almeida, and Aspenstrom 2013) as well as cellular migration (Li et al. 2015; Komatsu and Ikebe 2014), that may play unfavorably within the context of cancer metastasis. Invasion data were not available for the Toil Recomputed Compendium; however, an examination of *DAPK3* expression versus invasion for the ‘TCGA Colon Cancer’ dataset via Xena does reveal correlation between *DAPK3* expression and metastatic invasion occurrence. In this case, incidents of venous, lymphatic and perineural invasions were paradoxically associated with higher *DAPK3* expression (Figure 9C).

**Figure 8.**
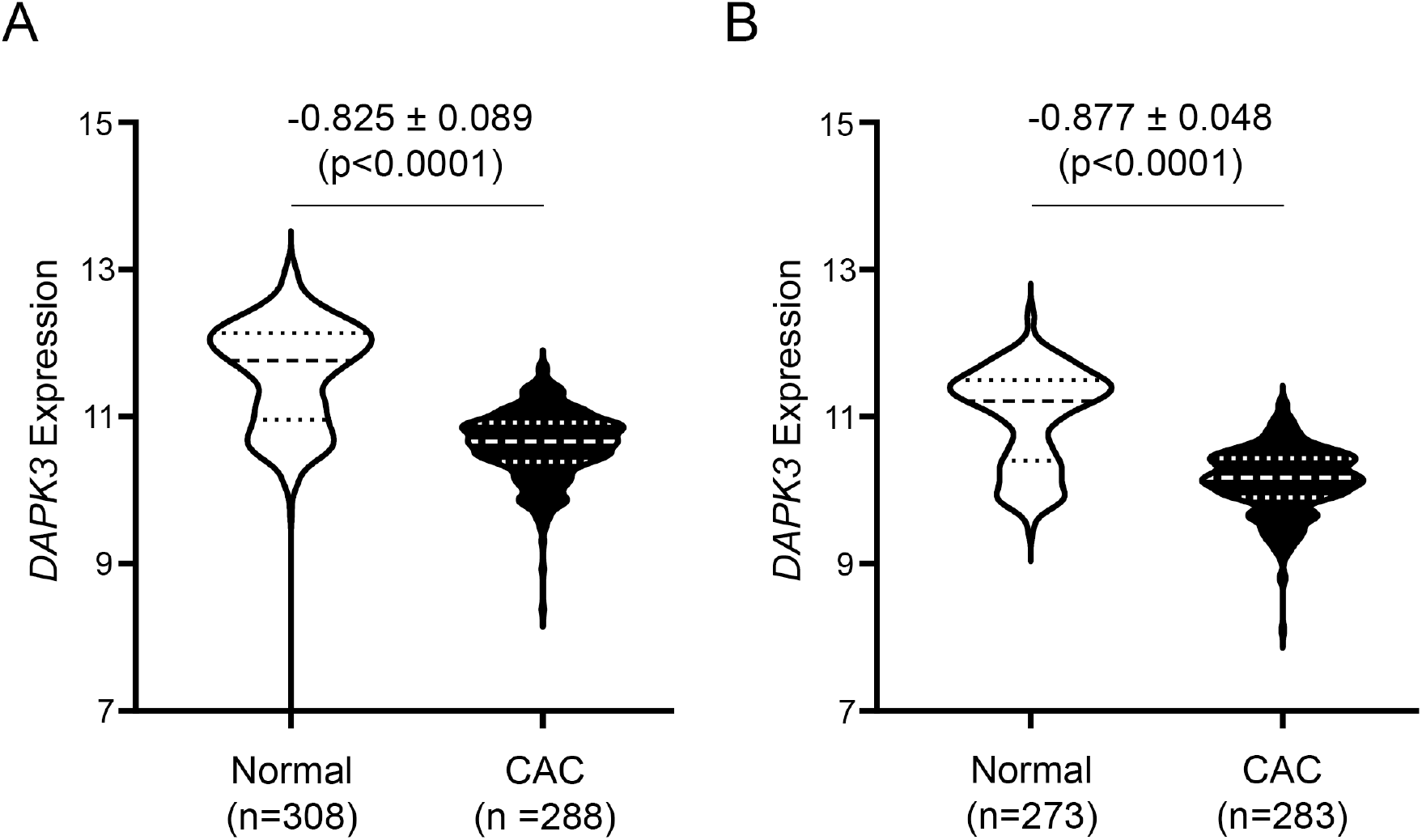
*DAPK3* expression in colon of healthy normal and primary colonic adenocarcinoma tumors. The expression of *DAPK3* in healthy colon (GTEx) and colon adenocarcinoma primary tumor (TCGA) was examined in UCSC Toil RNA-Seq Recompute Compendium (**A**) and MSKCC unified (**B**) datasets. Statistical significance was identified by unpaired Student’s t-test with Welch’s correction.

**Figure 9.**
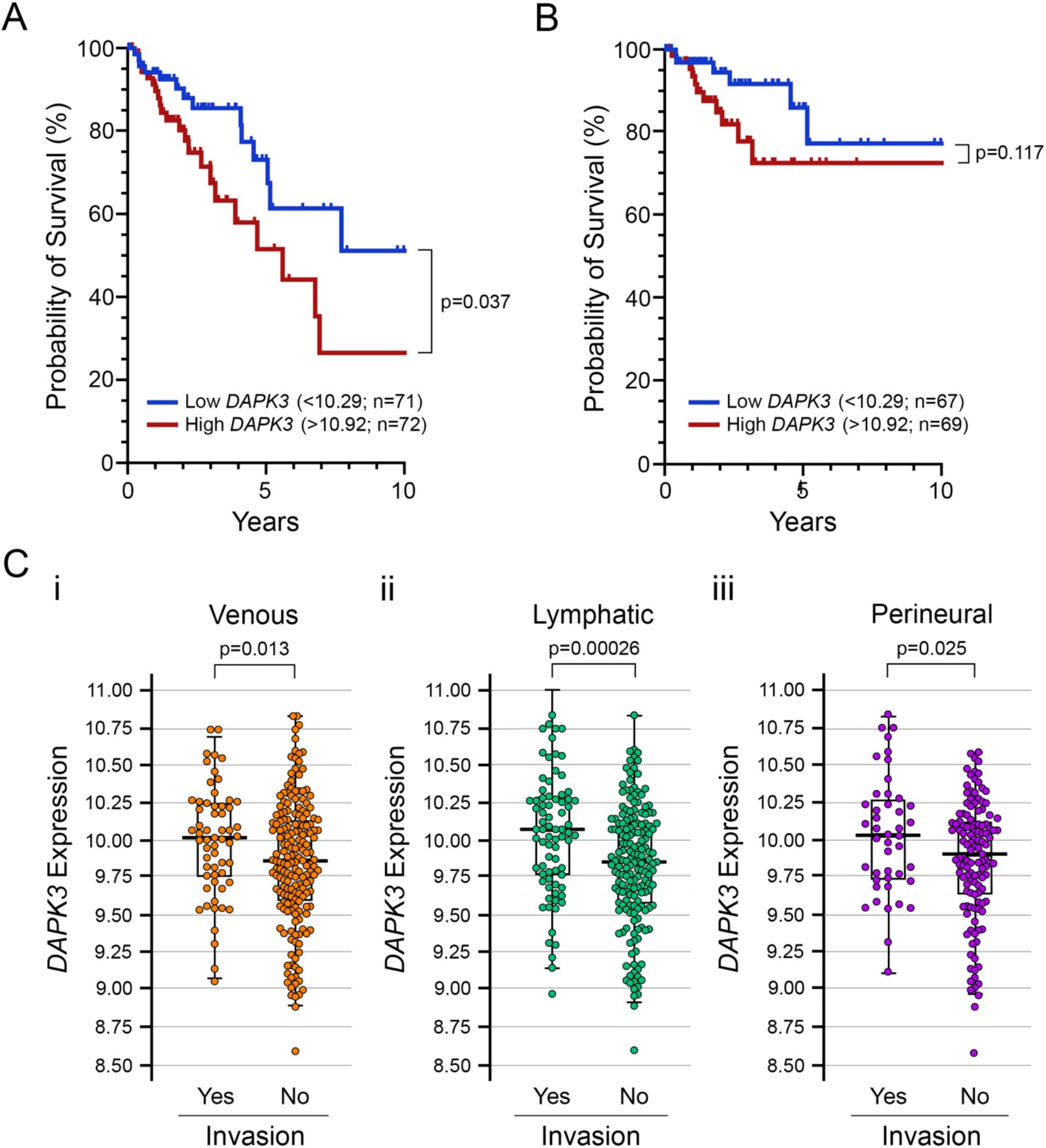
High *DAPK3* expression is associated with poor overall survival of patients with colon adenocarcinoma. Overall survival data (**A**) and disease specific survival data (**B**) were retrieved as part of the UCSC Toil Recompute dataset and subjected to Kaplan-Meier analyses. In (**C**), a comparison of *DAPK3* expression in colon adenocarcinoma patients with or without incidents of venous (i), lymphatic (ii) or perineural (iii) invasion was completed using data retrieved from the Xena TCGA data hub (https://tcga.xenahubs.net). Statistical significance was identified by unpaired Student’s t-test with Welch’s correction.

Genes that are functionally related tend to have similar expression profile; therefore, differential gene correlation analysis (DGCA) comparing expression correlation in normal and disease samples can illuminate *DAPK3*-dependent biological processes and/or molecular pathways that may be distinctly involved between the two states. DGCA was used to compare gene expression correlation in normal and colon adenocarcinoma disease samples, and the DGCA outcomes were sorted into nine possible categories (Figure 10). Of the 8,528 protein-coding genes surveyed, 8,222 (93%) were found to be differentially correlated with *DAPK3* between healthy controls and colon cancer conditions. When compared against the GTEx normal healthy population, *DAPK3* lost correlation with 46% of the genes surveyed and gained correlation with 47%. Finally, 6% of the surveyed genes had no significant correlation in either condition (healthy colon or colon adenocarcinoma) or had non-significant differential correlation. GO enrichment analysis was used to identify the biological processes and/or molecular pathways wherein differential correlation of *DAPK3* was implicated (Figure 11). *DAPK3* differential correlations changed most within the biological process of “Organic acid catabolic process; GO:0016054”, “Cell projection assembly; GO:0030031”, and “Muscle organ development; GO:0007517”, and the molecular pathways of “Endoribonuclease activity; GO:0004521” and “Extracellular matrix structural constituent; GO:0005201”.

**Figure 10.**
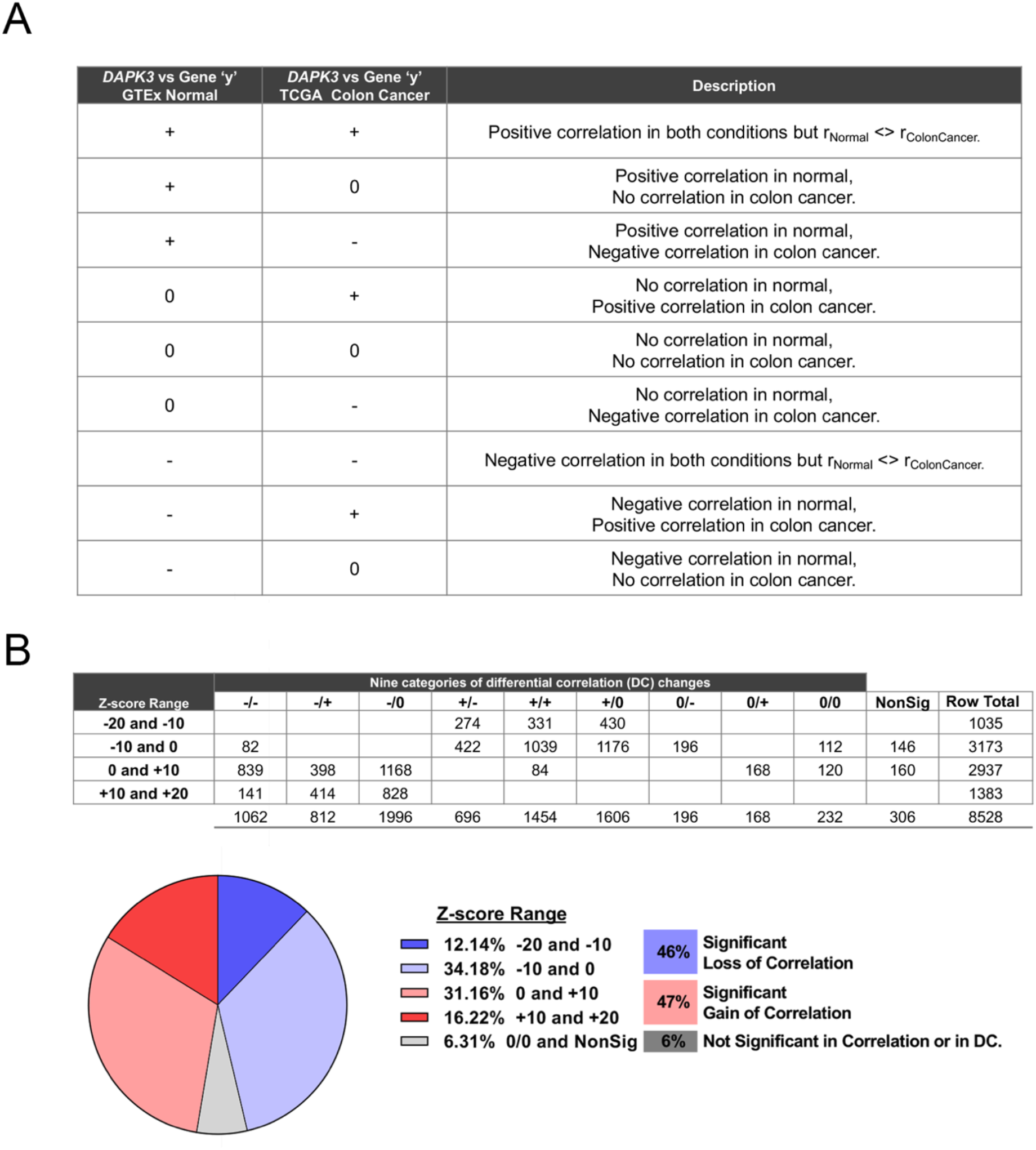
Differential gene correlation analysis (DGCA). (**A**), The DGCA analysis revealed nine categories of differential correlation changes for *DAPK3* in healthy colon and colon adenocarcinoma. (**B**), Breakdown of genes that displayed differentially correlation with *DAPK3* between healthy normal colon vs. CAC. Values represent gene counts.

**Figure 11.**
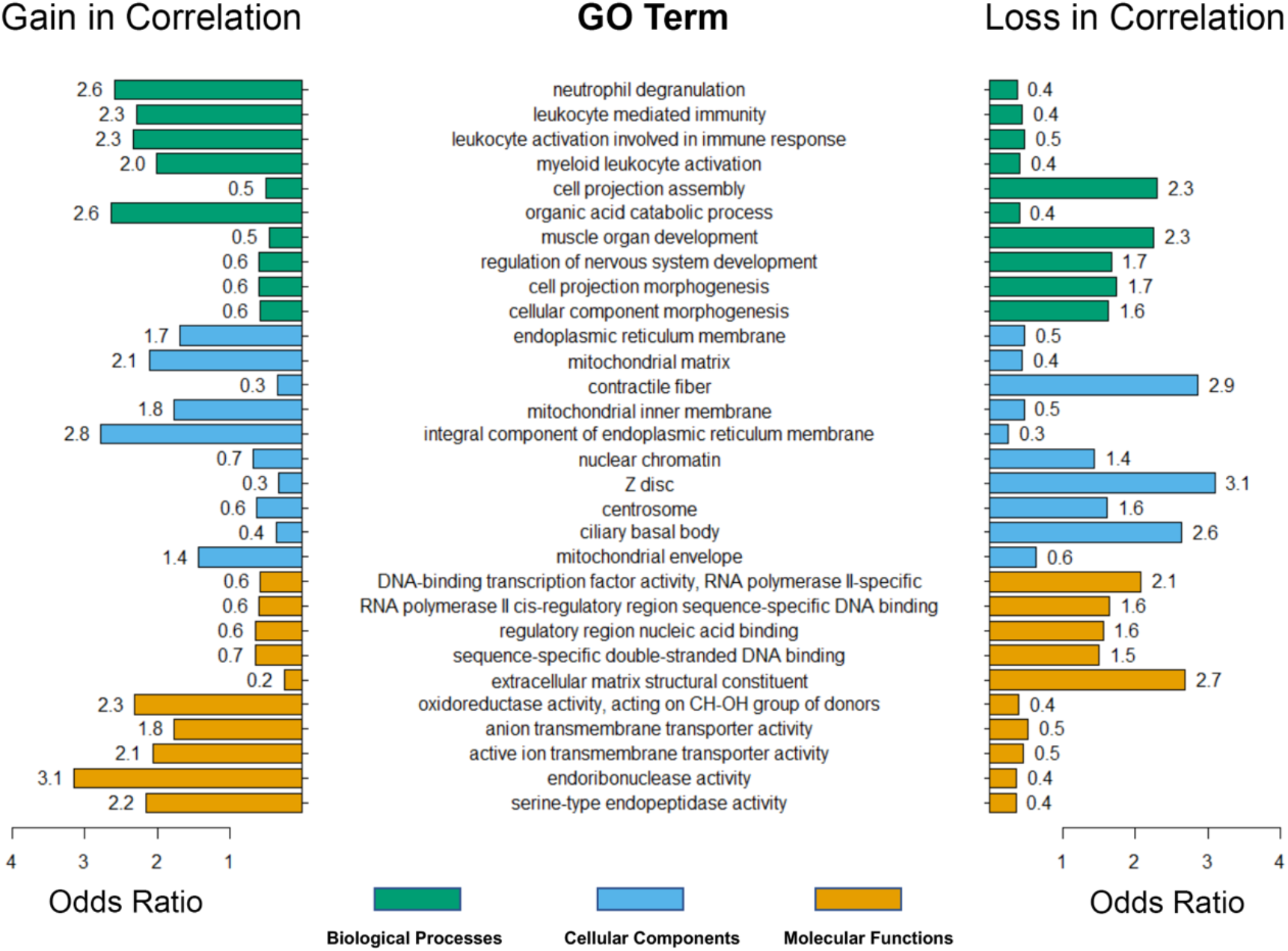
Differential gene correlation analysis (DGCA) of *DAPK3*-dependent biological processes. DGCA R was used to compare gene expression correlation in healthy and colon adenocarcinoma. GO enrichment analysis was completed using |minSize| = 30, |p-value cutoff| = 0.01, and |filterSigThresh| = 0.01.

## DISCUSSION

Several patient and disease characteristics are known to affect survival of patients with colon cancer, including age, sex, primary tumor location, tumor grade, and lymph node involvement (Dienstmann et al. 2019; Weiser et al. 2011). In addition, LVI and PNI are histopathological features associated with higher-risk colon cancer (Skancke et al. 2019; Mutabdzic et al. 2019; Cienfuegos et al. 2017; Liebig et al. 2009). However, the molecular biomarkers that distinguish between normal and invasive colon cancer are not well described. In this study, we employed the weighted gene co-expression network analysis (WGCNA) method to evaluate the system-level functionality of genes with clinical invasion features of colon adenocarcinoma.

The power of WGCNA lies in its ability to reveal gene modules (i.e., gene co-expression networks) and identify the central players (i.e., hub genes) within modules that are usually related to biological functions (Langfelder and Horvath 2008; Zhang and Horvath 2005; Li et al. 2018). RNA-Seq data and clinical information obtained from the TCGA database were included in the final WGCNA to identify robust co-expression modules associated with tumor invasion characteristics. A total of 24 distinct gene co-expression modules that included 9,750 protein coding genes were identified from primary tumors of colon cancer patients. After examining the associations between the modules and clinical invasion traits, *darkred* was identified to be a clinically significant gene module that required further interrogation. Significantly the module was linked with positive gene correlation towards lymphatic and venous invasion as well pathological stage. The 151 genes within this module were considered to possess functional associations with each other and, thus, could potentially identify important molecular pathways and hub genes that could advance understanding, detection and/or treatment opportunities (Langfelder and Horvath 2008).

Functional annotation of the *darkred* module revealed a core set of genes were involved in the “Actomyosin contractile ring organization” and the “Actin cytoskeleton pathway”. Moreover, the differential correlations identified for *DAPK3* with GO terms “Cell projection assembly” and “Extracellular matrix structural constituent” are of particular interest. These terms are associated with the molecular dynamics of cell migration and adhesion, which are particularly important to the process of metastasis. The cytoskeleton, especially the contractile tension generated by actomyosin, is also inherent to the pathogenesis and metastasis of tumors. Indeed, there is growing appreciation that a wide range of cellular activities that are highly relevant to tumorigenesis are dependent on the composition and organization of the cytoskeletal architecture, including tumor cell migration and invasion (Fletcher and Mullins 2010; Vasiliev 2004), epithelial-mesenchymal transformation (Rana et al. 2018; Lamouille, Xu, and Derynck 2014), nuclear dysmorphia and genome stability (Takaki et al. 2017; Irianto et al. 2016; Chow, Factor, and Ullman 2012), and tumor cell survival under hemodynamic shear stress (Xin et al. 2019). Furthermore, genes in the *darkred* module were also enriched in processes related to “Positive regulation of apoptotic signaling pathway” and “Regulation of mitochondrial membrane permeability involved in apoptotic process”. It is well known that the attenuation of apoptotic signaling is a hallmark of tumor biology (Zhang and Yu 2013; Yang et al. 2009), and a plethora of alterations have been revealed in key apoptotic pathways that increase tumor cell survival and reduce the efficacy of chemotherapy. Enhanced invasiveness is also observed in cancer cells that experience failed apoptosis (Berthenet et al. 2020) or incomplete mitochondrial outer membrane permeabilization (Ichim and Tait 2016).

When the genes in the *darkred* module were mapped using STRING, *DAPK3* was identified as a pseudohub gene. However, *RHOT2* (mitochondrial Rho GTPase 2; also known as *MIRO2*) was the true core hub based on node degree when the (i) differential expression or (ii) Kaplan-Meier overall survival estimate was disregarded. RHOT2 is a mitochondrial GTPase involved in mitochondrial trafficking and serves as a docking site for parkin (PRKN)-mediated mitophagy. While the RhoA GTPase family is closely associated with cancer progression, there are few studies on the role of the RHOT2 protein in tumor cell movement. In one published study, however, mitochondrial trafficking to the cortical cytoskeleton and tumor cell invasion were suppressed when siRNA-mediated silencing of Miro2 in LN229 glioma cells was used under conditions of cellular stress (Caino et al. 2016).

The *RHOT2* network also included *ENKD1, ARRDC1, NOL3, DAPK3, ANKRD13D* and *DEF8*. Each of these genes was differentially expressed in colon adenocarcinoma and was correlated with patient prognosis for overall survival analysis. The ANKRD13D protein possesses a ubiquitin-interacting motif (UIM) and forms a complex with other ANKRD13 family members to bind the Lys63-linked ubiquitin chains appended to epidermal growth factor receptor (EGFR) to regulate the internalization of ligand-activated EGFR (Tanno et al. 2012; Mattioni et al. 2020). *ANKRD13* has also been used in a panel of DNA methylation markers to identify colorectal cancer (CRC) in cell-free DNA samples obtained from patients (Cho et al. 2020). The ARDDC1 protein controls the generation of endosomal microvesicles that bud from the plasma membrane (Nabhan et al. 2012), the type of protein cargo (Anand et al. 2018), and, consequently, signal transduction by receptors subjected to endosomal sorting mechanisms. Other studies suggest that ARRDC1 may act as tumor suppressor by modulating the levels of Yes-associated protein (YAP) 1 and other membrane receptors (Xiao et al. 2018). NOL3 is an RNA-binding protein that has been implicated in tumorigenesis and metastasis. NOL3 was associated with worse prognosis in CRC and was overexpressed in CRC patients with lymph node metastasis (Zhang et al. 2020) (Toth et al. 2016). Finally, the gene products of *ENKD1* and *DEF8* are poorly described in the literature, so it is unclear how these gene products may act to impact upon tumor cells.

The emergence of *DAPK3* as a pseudohub gene in the *darkred* network with differential expression and significant linkage to survival outcome was particularly interesting. DAPK3 (also known as zipper-interacting protein kinase, ZIPK) is a serine/threonine protein kinase that acts as a molecular switch for a multitude of cellular processes, including the induction of apoptotic and autophagic cell death, cell proliferation, actomyosin contraction and cellular migration (Gozuacik and Kimchi 2006; Shiloh, Bialik, and Kimchi 2014; Usui, Okada, and Yamawaki 2014). The pro-apoptotic influence of DAPK3 is aligned with its association with promyelocytic leukemia (PML) oncogenic domains, nuclear structures implicated in transcription of regulation of apoptotic factors (Kawai, Akira, and Reed 2003). A marked drop in basal apoptosis is observed at the polyp-to-adenoma stage of colon cancer, suggesting that resistance to apoptotic programming is acquired early in tumorigenesis (Termuhlen et al. 2002). The reduced expression of *DAPK3*, as a tumor suppressing kinase, could provide a means for metastatic colon cancer cells to exhibit resistance to particular pathways of apoptosis. In addition, DAPK is associated with autophagic cell death; DAPK3 interaction with autophagy-related (ATG)-1 protein kinase allows for regulation of autophagosome formation via actomyosin activation (Tang et al. 2011). DAPK3 was also found to phosphorylate and increase the activity of Unc-51 like autophagy activating kinase (ULK1; the mammalian homolog of ATG1) and thereby drive autophagy induction (Li et al. 2020). Further to this finding, the co-expression of *ULK1* and *DAPK3* was correlated with favorable survival outcomes in gastric cancer patients (Li et al. 2020). DAPK3 promotes actin reorganization and actomyosin contraction by controlling the levels of myosin regulatory light chain phosphorylation (Komatsu and Ikebe 2004; Moffat et al. 2011) which has a broad range of cellular impacts, including cell migration, focal adhesion regulation, stress fiber bundling, and autophagic cell death. In addition, DAPK3 can attenuate myosin light chain phosphatase activity (Borman et al. 2002; MacDonald et al. 2001), and the loss of myosin phosphatase was further linked to cancer cell nuclear dysmorphia and genome instability (Takaki et al. 2017).

Although *DAPK*3 displays tumor suppressor properties (Li et al. 2015; Kocher, White, and Piwnica-Worms 2015; Bi et al. 2009; Das et al. 2016; Brognard et al. 2011; Li et al. 2020), it also plays an oncogenic role with regulation of transcriptional and translational programs that are tightly connected to cell survival, proliferation, and growth. In this regard, DAPK3 promotes epithelial-mesenchymal transition (Li et al. 2015; Kake et al. 2017). DAPK3 also provides transcriptional regulation of canonical Wnt/β-catenin signaling through an interaction with Nemo-like kinase to drive enhanced proliferation of colon carcinoma cells (Togi et al. 2011). Additionally, DAPK3 can directly enhance the transcriptional activity of both STAT3 and the DNA-binding androgen receptor (Sato et al. 2006; Felten et al. 2013). As previously hypothesized by Kake and colleagues (Kake et al. 2017), DAPK3 may change from a tumor suppressor into a tumor promoter during the course of tumor progression. In the present study, we show that *DAPK3* expression is reduced in samples obtained from colon cancer patients; however, there is significant correlation for *DAPK3* association with LVI and PNI properties and worse survival probability. Further investigations will be required to solve the incongruity observed with respect to the pro-apoptotic role of DAPK3 in tumor suppression versus its proliferative influence on tumorigenesis.

## Data Availability

RNA sequencing data (RNA-Seq) for the Genotype Tissue Expression project (GTEx; https://gtexportal.org/home/) and The Cancer Genome Atlas (TCGA; https://portal.gdc.cancer.gov) were retrieved from the Xena Toil RNA-Seq Recompute Compendium (https://toil.xenahubs.net). TCGA survival data and COAD clinicalMatrix information of colon cancer patients were retrieved from the Xena TCGA data hub (https://tcga.xenahubs.net).

https://toil.xenahubs.net

## ACKNOWLEDGEMENTS

This work was supported by a research grant from the Canadian Institutes of Health Research (MOP#97931 to J.A.M.). H.-M.C. was recipient of a CIHR Fredrick Banting and Charles Best Canada Graduate Scholarship.

## AUTHOR CONTRIBUTIONS

H.-M.C. completed the data analysis. H.-M.C. and J.A.M. prepared figures. J.A.M. conceived and coordinated the study, wrote the manuscript, supervised trainees and provided intellectual contributions to the project. All authors reviewed the results and approved the final version of the manuscript.

## AUTHORS’ COMPETING INTERESTS STATEMENT

J.A.M. is cofounder and has an equity position in Arch Biopartners Inc. All other authors declare no conflicts of interest.

## Notes

### Competing Interest Statement

J.A. MacDonald is cofounder and has an equity position in Arch Biopartners Inc. All other authors declare no conflicts of interest.

### Author Declarations

No IRB and/or ethics committee approvals were required.

